# Sex-Specific Genetic Drivers of Memory, Executive Functioning, and Language Performance in Older Adults

**DOI:** 10.1101/2025.05.26.25328369

**Authors:** Jaclyn M. Eissman, Alexandra N. Regelson, Skylar Walters, Derek B. Archer, Alaina Durant, Shubhabrata Mukherjee, Michael L. Lee, Seo-Eun Choi, Phoebe Scollard, Emily H. Trittschuh, Jesse Mez, Moonil Kang, William S. Bush, Brian W. Kunkle, Adam C. Naj, Katherine A. Gifford, Murat Bilgel, Amanda B. Kuzma, The Alzheimer’s Disease Neuroimaging Initiative (ADNI), The Alzheimer’s Disease Genetics Consortium (ADGC), The Alzheimer’s Disease Sequencing Project (ADSP), Michael L. Cuccaro, Carlos Cruchaga, Margaret A. Pericak-Vance, Lindsay A. Farrer, Li-San Wang, Gerard D. Schellenberg, Badri N. Vardarajan, Richard Mayeux, Jonathan L. Haines, Angela L. Jefferson, Walter A. Kukull, C. Dirk Keene, Andrew J. Saykin, Paul M. Thompson, Eden R. Martin, Marilyn S. Albert, Sterling C. Johnson, Corinne D. Engelman, Luigi Ferrucci, David A. Bennett, Lisa L. Barnes, Julie A. Schneider, Susan M. Resnick, Reisa A. Sperling, Paul K. Crane, Timothy J. Hohman, Logan Dumitrescu

**Affiliations:** Vanderbilt Memory & Alzheimer’s Center, Vanderbilt University Medical Center, Nashville, TN, 37203, USA; Vanderbilt Genetics Institute, Vanderbilt University Medical Center, Nashville, TN, 37232, USA; Department of Anatomy and Neurobiology, Boston University Chobanian & Avedisian School of Medicine, Boston, MA, 02118, USA; Department of Medicine, University of Washington, Seattle, WA, 98195, USA; Department of Psychiatry and Behavioral Sciences, University of Washington School of Medicine, Seattle, WA, 98195, USA; VA Puget Sound Health Care System, GRECC, Seattle, WA, 98108, USA; Department of Neurology, Boston University Chobanian & Avedisian School of Medicine, Boston, MA, 02118, USA; Cleveland Institute for Computational Biology, Department of Population and Quantitative Health Sciences, School of Medicine, Case Western Reserve University, Cleveland, OH, 44106, USA; John P. Hussman Institute for Human Genomics, University of Miami Miller School of Medicine, Miami, FL, 33136, USA; Department of Biostatistics, Epidemiology, and Informatics, University of Pennsylvania Perelman School of Medicine, Philadelphia, PA, 19104, USA; Penn Neurodegeneration Genomics Center, Department of Pathology and Laboratory Medicine, University of Pennsylvania Perelman School of Medicine, Philadelphia, PA, 19104, USA; Laboratory of Behavioral Neuroscience National Institute on Aging, Baltimore, MD, 21224, USA; Department of Psychiatry, Washington University School of Medicine, St. Louis, MO, 63110, USA; NeuroGenomics and Informatics Center, Washington University School of Medicine, St. Louis, MO, 63108, USA; Department of Biostatistics, Boston University School of Public Health, Boston, MA, 02118, USA; Department of Medicine Biomedical Genetics, Boston University Chobanian & Avedisian School of Medicine, Boston, MA, 02118, USA; Taub Institute for Research on Alzheimer’s Disease and the Aging Brain, Columbia University, New York, NY, 10032, USA; Gertrude H. Sergievsky Center, Columbia University, New York, NY, 10032, USA; Vagelos College of Physicians and Surgeons, Columbia University, New York, NY, 10032, USA; Department of Epidemiology, School of Public Health, University of Washington, Seattle, WA, 98195, USA; Department of Laboratory Medicine and Pathology, University of Washington, Seattle, WA, 98195, USA; Department of Radiology and Imaging Services, Indiana University School of Medicine, Indianapolis, IN, 46202, USA; Department of Medical and Molecular Genetics, School of Medicine, Indiana University, Indianapolis, IN, 46202, USA; Indiana Alzheimer’s Disease Research Center, Indiana University School of Medicine, Indianapolis, IN, 46202, USA; Imaging Genetics Center, Stevens Neuroimaging & Informatics Institute, Keck School of Medicine, University of Southern California, Los Angeles, CA, 90033, USA; Department of Neurology, Johns Hopkins University School of Medicine, Baltimore, MD, 21205, USA; Alzheimer’s Disease Research Center, University of Wisconsin School of Medicine and Public Health, Madison, WI, 53792, USA; Department of Population Health Sciences, University of Wisconsin School of Medicine and Public Health, Madison, WI, 53792, USA; Longitudinal Studies Section, Translational Gerontology Branch, National Institute on Aging, Baltimore, MD, USA; Rush Alzheimer’s Disease Center, Rush University Medical Center, Chicago, IL, 60612, USA; Department of Neurology, Harvard Medical School, Boston & Brigham and Women’s Hospital, Boston, MA, 02115, USA; Alzheimer’s Disease Research Center, Massachusetts General Hospital, Charlestown, MA, 02129, USA

## Abstract

We previously identified sex-specific genetic loci associated with memory performance, a strong Alzheimer’s disease (AD) endophenotype. Here, we expand on this work by conducting sex-specific, cross-ancestral, genome-wide meta-analyses of three cognitive domains (memory, executive functioning, and language) in 33,918 older adults (57% female; 41% cognitively impaired; mean age=73 years) from 10 aging and AD cohorts. All three domains were comparably heritable across sexes. Genome-wide meta-analyses identified three novel loci: a female-specific language decline-associated locus, *VRK2* (rs13387871), which is a published candidate for neuropsychiatric traits involving language ability; a male-specific memory decline-associated locus among cognitive impaired, *DCHS2* (rs12501200), which is a published candidate gene for AD age-at-onset; and a sex-interaction with baseline executive functioning, *AGA* (rs1380012), among cognitive impaired. We additionally provide evidence for shared genetic architecture between lifetime estrogen exposure and AD-related cognitive decline. Overall, we identified sex-specific variants, genes, and pathways relating to three cognitive domains among older adults.

## Main

Women are disproportionality affected by Alzheimer’s disease (AD). Two-thirds of patients with AD are women^1^ and lifetime risk of AD is two-fold greater in women compared to men^2,3^. Sex differences may, in part, be due to greater longevity in women^4,5^. However, evidence shows differences in the clinical presentation of AD by sex, including a greater neuropathological burden in women compared to men^6,7^, on average, often most pronounced in the later stages of AD neuropathological progression^6^. Moreover, there are notable sex differences in the clinical manifestation of disease, with women typically showing a faster rate of cognitive decline in the presence of enhanced AD neuropathological burden^7^ or enhanced biomarkers of AD neuropathology^8^. Thus, there is a biological basis for examining sex differences in relation to the AD neuropathological cascade, including its downstream consequences, like cognitive impairment.

Cognitive changes in AD tend to involve deficits across multiple domains, including memory, executive functioning, and language, with memory being the most extensively studied. In the absence of disease, in general, there is a clear female advantage in verbal memory performance^9,10^. Sundermann and colleagues (among others) have detailed a female verbal memory advantage in preclinical AD, with preclinical women showing better immediate and delayed recall among those with moderate amyloid burden^11^, hippocampal atrophy^12^, and brain hypometabolism^13^ as compared to preclinical men. Notably, these prior studies show an attenuation of the female advantage in those with AD, suggesting the possibility of female-specific cognitive reserve in preclinical stages, but a faster progression, compared to men, after dementia onset^11–13^. This sex difference in AD-related cognition extends beyond memory performance. Evidence shows that women typically have higher baseline executive functioning^14^ and language performance^15^, but demonstrate more rapid global cognitive decline^14^, possibly driven by the presence of AD neuropathology^8^. Taken together, this evidence may suggest sex differences in cognitive performance, including by cognitive domain, that may change throughout the course of disease.

To explore the genetic architecture of late-life cognitive decline, our group has derived harmonized cognitive scores as part of the Alzheimer’s Disease Sequencing Project Phenotype Harmonization Consortium (ADSP-PHC). The approach to harmonization uses item response theory to derive robust, co-calibrated composite measures of memory, executive functioning, and language that can be leveraged in genomic studies^16^. Our group has leveraged these data to characterize the genetic architecture, and notably, the sex differences in genetic architecture, of both memory decline^17,18^ and cognitive resilience^19,20^ (i.e., better-than-expected cognition given amyloid pathology burden). These published works demonstrate that we can identify sex-specific loci associated with AD-related traits that do not appear in the sex-agnostic versions of these GWAS^18,20^.

In this study, we sought to expand on our previous work by conducting a cross-ancestral, genetic analysis of three domains of cognition, both with baseline measures and with measures of longitudinal cognitive decline. While our prior study evaluated genetic drivers of memory performance in 4 cohorts of aging and AD, this present study includes 10 cohorts and nearly 34,000 individuals with harmonized cognitive scores across memory, executive functioning, and language domains. We hypothesized that we would identify genetic drivers across the three cognitive domains that would differ by sex, which will aid in our understanding of sex-specific cognitive changes in preclinical and clinical AD.

## Results

Participant characteristics are presented in **Table 1**. Detailed information about each cohort included in this study can be found in **Supplementary Methods I**.

**Table 1.** Participant Characteristics.

|  | **Non-Hispanic White**<br>**(N=30,432)** |  | **Non-Hispanic Black**<br>**(N=3,486)** |  | **Cross-Ancestry**<br>**(N=33,918)** |  |
| --- | --- | --- | --- | --- | --- | --- |
|  | **<u>Males</u>** | **<u>Females</u>** | **<u>Males</u>** | **<u>Females</u>** | **<u>Males</u>** | **<u>Females</u>** |
| N (%) | 13,617 (44.7%) | 16,815 (55.3%) | 922 (26.4%) | 2,564 (73.6%) | 14,539 (42.9%) | 19,379 (57.1%) |
| **Baseline Characteristics** |  |  |  |  |  |  |
| Age (years), mean +/- SD | 73.1 (8.5) | 73.0 (9.0) | 71.1 (8.9) | 72.3 (8.6) | 73.0 (8.5) | 72.9 (8.9) |
| Education (years), mean +/- SD | 16.4 (3.1) | 15.5 (2.9) | 14.5 (3.4) | 14.6 (3.1) | 16.3 (3.1) | 15.4 (2.9) |
| Cognitively unimpaired, N (%) | 7,112 (52.2%) | 10,957 (65.2%) | 438 (47.5%) | 1,470 (57.3%) | 7,550 (51.9%) | 12,427 (64.1) |
| **Baseline Cognition** |  |  |  |  |  |  |
| Memory score, mean +/- SD | 0.2 (0.8) | 0.4 (0.9) | 0.0 (0.8) | 0.2 (0.8) | 0.2 (0.8) | 0.4 (0.9) |
| Executive functioning score, mean +/- SD | 0.1 (0.8) | 0.2 (0.8) | -0.3 (0.8) | -0.1 (0.7) | 0.1 (0.8) | 0.2 (0.8) |
| Language score, mean +/- SD | 0.3 (0.8) | 0.4 (0.8) | 0.1 (0.7) | 0.2 (0.7) | 0.2 (0.8) | 0.4 (0.8) |
| **Longitudinal Characteristics** |  |  |  |  |  |  |
| Has longitudinal data available, N (%) | 10,172 (74.7%) | 12,385 (73.7%) | 726 (78.7%) | 2,059 (80.3%) | 10,898 (75.6%) | 14,444 (74.5%) |
| Number of visits (mean +/- SD) | 5.5 (3.6) | 5.9 (3.9) | 5.4 (3.5) | 5.9 (4.0) | 5.5 (3.6) | 5.9 (3.9) |
| Years of follow-up (mean +/- SD) | 6.3 (5.6) | 6.9 (5.4) | 6.3 (5.5) | 6.6 (5.3) | 6.3 (5.6) | 6.8 (5.4) |
| **Longitudinal Cognitive Decline** |  |  |  |  |  |  |
| Memory decline slope (mean +/- SD) | -0.1 (0.1) | -0.1 (0.1) | -0.1 (0.1) | -0.1 (0.1) | -0.1 (0.1) | -0.1 (0.1) |
| Executive functioning decline slope (mean +/- SD) | -0.1 (0.1) | -0.1 (0.1) | -0.1 (0.1) | -0.1 (0.1) | -0.1 (0.1) | -0.1 (0.1) |
| Language decline<br>slope (mean +/- SD) | -0.1 (0.1) | -0.1 (0.1) | -0.1 (0.1) | -0.1 (0.1) | -0.1 (0.1) | -0.1 (0.1) |
| ***APOE* Genotype Information** |  |  |  |  |  |  |
| *APOE* ε4 carrier,<br>N (%) | 5,094 (37.4%) | 6,161 (36.6%) | 436 (47.3%) | 1,097 (42.8%) | 5,530 (38.0%) | 7,258 (37.5%) |
| *APOE* ε2 carrier,<br>N (%) | 1,657 (12.2%) | 2,048 (12.2%) | 183 (19.8%) | 480 (18.7%) | 1,840 (12.7%) | 2,528 (13.0%) |

### Heritability of Cognitive Endophenotypes by Sex

We calculated SNP-based heritability estimates in each sex for each of the 6 cognitive endophenotypes (**Table 2**), and then we tested to see if estimates significantly differed by sex, leveraging a previously published approach^18,21^. Estimates were calculated separately in non-Hispanic white (NHW) and non-Hispanic black (NHB) individuals. However, estimates were unstable in NHB, likely due to sample size restrictions. Therefore, only estimates among NHW individuals are reported in **Table 2**. SNP-based heritability estimates were generally higher in men (h^2^: 0.14-0.21) than in women (h^2^: 0.11-0.17) across the 3 domains for both baseline and longitudinal cognitive measures, but heritability estimates did not significantly differ between sexes for any of the cognitive domains (**Table 2**).

**Table 2.** Sex-Specific SNP-Based Heritability Estimation.

| **Baseline Cognition** |  |  |  |  |  |  |
| --- | --- | --- | --- | --- | --- | --- |
|  | **Males** |  | **Females** |  | **Sex Differences Test** |  |
|  | **V(G)/Vp (SE)** | **P-value** | **V(G)/Vp (SE)** | **P-value** | **Z-score** | **P-value** |
| Baseline Memory | 0.14 (0.03) | $5.51 \times 10^{-12}$ | 0.14 (0.02) | $1.77 \times 10^{-14}$ | 0.00 | 1.00 |
| Baseline Executive Functioning | 0.21 (0.03) | $<<<0.01^*$ | 0.17 (0.02) | $<<<0.01^*$ | -1.11 | 0.27 |
| Baseline Language | 0.15 (0.03) | $2.97 \times 10^{-10}$ | 0.11 (0.02) | $1.13 \times 10^{-7}$ | -1.11 | 0.27 |
| **Cognitive Decline** |  |  |  |  |  |  |
|  | **Males** |  | **Females** |  | **Sex Differences Test** |  |
|  | **V(G)/Vp (SE)** | **P-value** | **V(G)/Vp (SE)** | **P-value** | **Z-score** | **P-value** |
| Memory Decline | 0.17 (0.03) | $3.32 \times 10^{-10}$ | 0.12 (0.03) | $9.56 \times 10^{-8}$ | -1.18 | 0.24 |
| Executive Functioning Decline | 0.14 (0.03) | $6.65 \times 10^{-7}$ | 0.11 (0.03) | $4.87 \times 10^{-7}$ | -0.71 | 0.48 |
| Language Decline | 0.16 (0.03) | $2.52 \times 10^{-8}$ | 0.11 (0.03) | $3.62 \times 10^{-7}$ | -1.18 | 0.24 |

Note: Reported values are V(G)/Vp (Standard Error – SE); \*P-value estimated as 0 through GCTA

### Cross-Ancestry Meta-Analysis of Cognitive Endophenotypes by Sex

We conducted sex-stratified and sex-interaction genome-wide meta-analyses in each ancestry group (i.e., based on both self-report as well as genetic similarity to 1000G EUR and AFR reference populations) and then meta-analyzed across groups. We also performed secondary analyses stratified by clinical diagnosis: cognitively unimpaired (CU) and cognitively impaired (CI) individuals. All loci that reached genome-wide suggestive significance (p<1×10^−5^) are presented in **Supplementary Tables 1-6**. We also leveraged sex-stratified GWAS summary statistics of cognition-related traits from the Framingham Heart Study (FHS^22^) and the UK Biobank (UKBB^23^) for replication. Please see **Methods** for details on all models. Below, we will be presenting all genome-wide significant (p<5×10^−8^) loci beyond the well-known *APOE* locus.

We identified a genome-wide significant locus on chromosome 2 (**Figure 1**; **Supplementary Figure 1**) associated with slower language decline among females but not among males (index SNP: rs13387871; MAF_both_sexes_=0.20; β_females_=2.67×10^−3^, p_females_=5.97×10^−9^; β_males_=-4.42×10^−4^, p_males_=0.44), and the sex-interaction (at rs13387871) nearly reached genome-wide suggestive significance (β_interaction_=2.88×10^−3^, p_interaction_=6.08×10^−5^), further suggesting that this locus may differ by sex. The chromosome 2 locus was more strongly driven by cross-ancestral CU females (β_females_=2.27×10^−3^, p_females_=5.01×10^−7^) as opposed to the CI females (β_females_=4.89×10^−3^, p_females_=1.29×10^−2^). Additionally, rs13387871 had a consistent magnitude and direction of effect across cognitive domains and was nominally significant for memory decline (β_females_=1.59×10^−3^, p_females_=6.32×10^−3^) and fell just short of nominal significance for executive functioning decline (β_females_=5.61×10^−4^, p_females_=0.06). When adjusting for educational attainment, the female-specific association between rs13387871 and language decline only attenuated slightly (β_females_=2.33×10^−3^, p_females_=1.58×10^−7^). This finding did not replicate in FHS or in UKBB.

**Figure 1.**
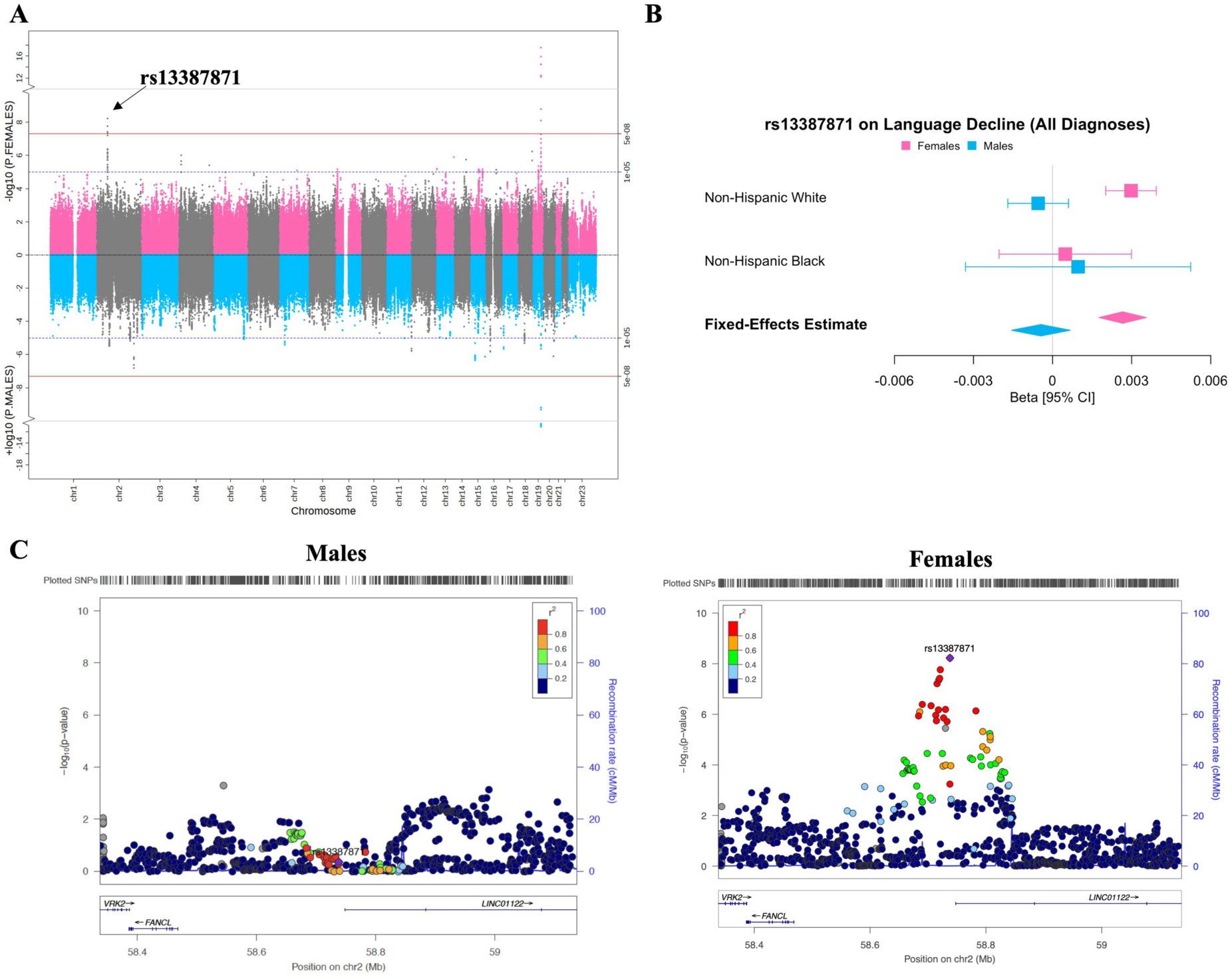
Genome-wide significant locus associated with female-specific language decline. Minor allele of genome-wide significant chromosome 2 locus, led by rs13387871, associated with rate of language decline among cross-ancestral females of all diagnoses. (**A**) Miami plot displays variant associations with language decline among females (top) in pink and males (bottom) in blue. (**B**) Forest plot of lead variant, rs13387871, displays 95% confidence intervals of fixed-effects meta-analysis estimates in each ancestry group and cross-ancestrally. (**C**) Regional plot displays the genomic region surrounding index variant, rs13387871, by sex.

Diagnosis subgroup meta-analyses identified 2 additional loci associated with cognition among CI individuals. First, we identified a genome-wide significant locus on chromosome 4 (**Figure 2**; **Supplementary Figure 2**) associated with faster memory decline among CI males but not among CI females (index SNP: rs12501200; MAF_both_sexes_=0.27; β_males_=-1.25×10^−2^, p_males_=3.21×10^−8^; β_females_=1.79×10^−3^, p_females_=0.43), and this locus had a sex-interaction (among CI individuals) with memory decline that nearly reached genome-wide suggestive significance (β_interaction_=1.40×10^−2^, p_interaction_=2.06×10^−5^). Additionally, rs12501200 had a consistent magnitude and direction of effect across both executive functioning decline (β_males_=-2.66×10^−3^, p_males_=0.07) and language decline (β_males_=-7.05×10^−3^, p_males_=5.99×10^−5^), nearly reaching genome-wide suggestive significance for language decline. When adjusting for educational attainment, the association with memory decline was slightly attenuated among the cross-ancestral sample of CI males (β_males_=-1.11×10^−2^, p_males_=1.65×10^−6^). We replicated the association with rs12501200 in a published male-specific GWAS from the UKBB^23^ of “subset inclusive logic from fluid intelligence,” a trait related to working memory (β_males_=-0.06, p_males_=7.94×10^−3^).

**Figure 2.**
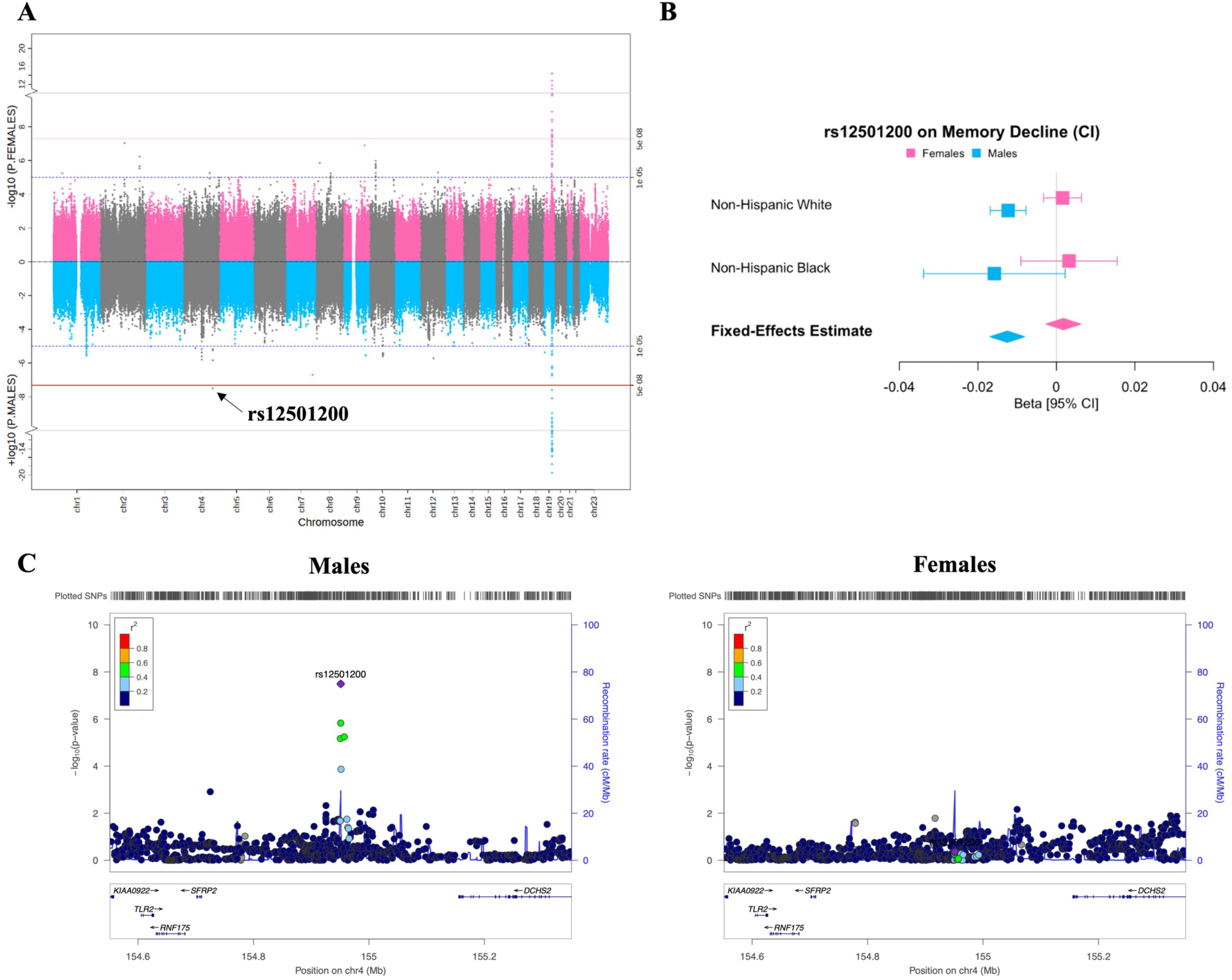
Genome-wide significant locus associated with memory decline among cognitively impaired males. Minor allele of genome-wide significant chromosome 4 locus, led by rs12501200, associated with rate of memory decline among cross-ancestral males with cognitive impairment. (**A**) Miami plot displays variant associations with memory decline among cognitively impaired females (top) in pink and cognitively impaired males (bottom) in blue. (**B**) Forest plot of lead variant, rs12501200, displays 95% confidence intervals of fixed-effects meta-analysis estimates in each ancestry group and cross-ancestrally. (**C**) Regional plot displays the genomic region surrounding index variant, rs12501200, by sex.

Second, we identified a genome-wide significant sex-interaction (**Figure 3**; **Supplementary Figure 3**) on chromosome 4 with baseline executive functioning among CI individuals (index SNP: rs1380012; MAF_both_sexes_=0.26; β_interaction_=0.13, p_interaction_=2.40×10^−8^), that was nominally significant and had opposing directions of effect among CI males and females (β_males_=-0.06, p_males_=2.76×10^−4^; β_females_=4.82×10^−2^, p_females_=2.15×10^−3^). The interaction was nonsignificant among the CU sample (β_interaction_=-9.50×10^−4^, p_interaction_=0.94). Additionally, rs1380012 had nominally significant sex-interactions (similar magnitude and direction of effect) across domains for both baseline memory (β_interaction_=0.08, p_interaction_=1.30×10^−3^) and baseline language (β_interaction_=0.05, p_interaction_=1.65×10^−2^). When adjusting for educational attainment, the sex-interaction with baseline executive functioning attenuated to nonsignificant (β_interaction_=0.31, p_interaction_=0.68). This finding did not replicate in FHS or in UKBB.

**Figure 3.**
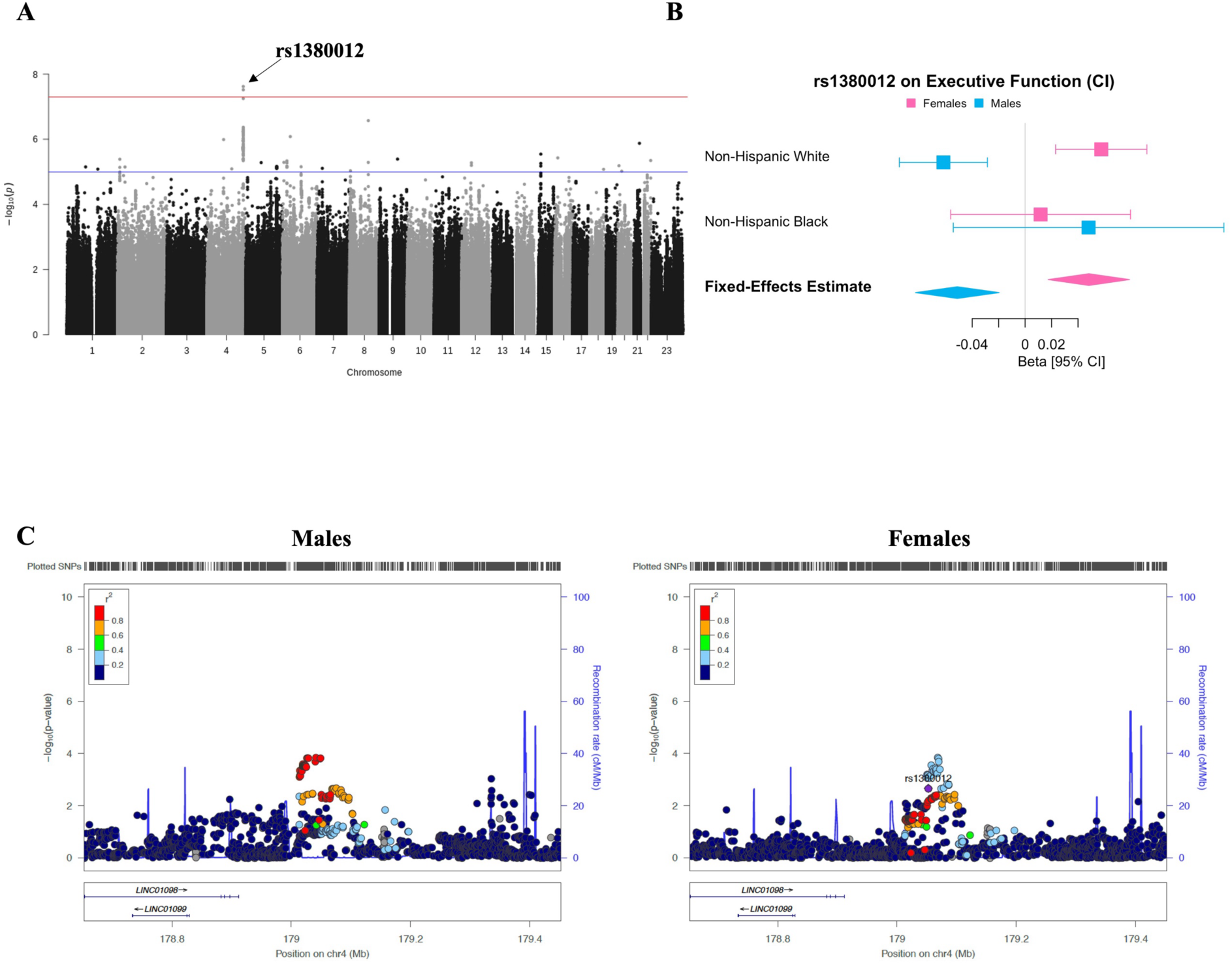
Genome-wide significant sex-interaction with baseline executive functioning among cognitively impaired individuals. Minor allele of genome-wide significant sex-interaction at chromosome 4 locus, led by rs1380012, associated with baseline executive functioning performance among cross-ancestral individuals with cognitive impairment. (**A**) Manhattan plot displays variant sex-interactions with baseline executive functioning among cognitively impaired individuals. (**B**) Forest plot of lead variant, rs1380012, displays 95% confidence intervals of fixed-effects meta-analysis estimates in each ancestry group and cross-ancestrally. (**C**) Regional plot displays the genomic region surrounding index variant, rs1380012, by sex.

### Candidate Gene Prioritization of Genome-Wide Significant Loci

We applied the following steps to nominate candidate genes associated with cognitive endophenotypes: 1) quantitative trait loci (QTL) analysis, for index SNPs of genome-wide significant loci (from the meta-analyses) leveraging public databases, 2) gene-level tests surviving genome-wide adjustment for multiple comparisons, leveraging the MAGMA^24^ software package, 3) query of genes from steps #1 and #2 in the AGORA AMP-AD knowledge portal, and 4) evaluation of known AD GWAS loci, with published sex-specificity, for associations with cognitive endophenotypes by sex. Our gene prioritization strategies resulted in the following candidate genes as having sex-specificity with respect to cognitive endophenotypes: *FANCL, VRK2, DCHS2, TRIM2, AGA*, *KCNA10, TNN, TYSND1, SAR1A, BIN1*, *PICALM*, *MS4A6A*, and *SLC9A7*. We will outline the biological evidence for each of these genes here.

We evaluated expression QTL (eQTL) data from public databases for the 3 genome-wide significant lead variants from the cross-ancestral meta-analyses: rs13387871 (female-specific language decline), rs12501200 (male-specific memory decline in CI), and rs1380012 (sex-interaction with executive functioning in CI). First, rs13387871 is an eQTL in CD8+ T-cells (β=-2.35×10^−1^, p=2.52×10^−2^) in blood for *FANCL* (an E3 ubiquitin-protein ligase in the Fanconi anemia pathway) and in monocytes (β=1.07×10^−1^, p=2.86×10^−2^) in blood for *VRK2* (a serine/threonine kinase involved with apoptosis). Additionally, rs13387871 is an eQTL for both *FANCL* (β=1.36×10^−1^, p=1.09×10^−2^) and *VRK2* (β=1.31×10^−1^, p=2.03×10^−2^) in serotonergic neurons. Second, the chromosome 4 locus index SNP rs12501200 is both an eQTL in B-cells (β=2.05×10^−1^, p=3.56×10^−2^) in blood and for inhibitory neurons in brain cortex (β=1.76×10^−1^, p=4.15×10^−3^) for *DCHS2* (encodes for proteins involved with cell adhesion). Additionally, rs12501200 is an eQTL in brain for progenitor cells (β=3.89×10^−2^, p=4.46×10^−2^), in brain cortex for microglia (β=-1.28×10^−1^, p=2.76×10^−2^), and for neurons (β=-5.53×10^−2^, p=2.00×10^−2^) all for *TRIM2* (E3 ubiquitin ligase activity in proteasome-mediated protein degradation). Third, the chromosome 4 locus with index SNP rs1380012 is an eQTL in blood for CD4+ T-cells (β=-1.13×10^−1^, p=4.88×10^−2^), monocytes (β=-3.30×10^−1^, p=2.56×10^−2^), and natural killer cells (β=-3.09×10^−1^, p=3.74×10^−2^), all for *AGA* (encodes for a protein involved with catabolism of N-linked glycoproteins).

Gene-level analyses (**Supplementary Tables 7-12**) conducted with MAGMA identified 4 additional candidate genes that survived genome-wide adjustment for multiple comparisons (false-discovery rate - FDR<0.05). First, we identified *KCNA10*, which had a significant sex-interaction with baseline executive functioning (among individuals of all diagnoses; P.FDR_interaction_=0.02) and showed nominal significance in both males (p_males_=8.52×10^−4^) and females (p_females_=1.32×10^−2^). In addition, we identified 3 genes, *TYSND1*, *SAR1A*, and *TNN*, that significantly associated with the rate of memory decline among CI males (P.FDR_males_<0.04). All 3 of these genes had at least a nominally significant (p<0.05) sex-interaction with memory decline (CI individuals), and the sex-interaction at *TNN* with memory decline additionally survived FDR correction (P.FDR_interaction_=0.03).

Next, we queried all genes from the prior 2 steps in the AGORA AMP-AD knowledge portal, and evidence is reported here if it aligns with the sex of which the locus or gene was originally identified in our variant- or gene-level analysis. First, *VRK2* is differentially expressed (log_2_ fold change – log_2_FC) in the dorsolateral prefrontal cortex (DLPFC) and parahippocampal gyrus (PHG) between female AD cases and controls (log_2_FC_DLPFC_=0.24, p_DLPFC_=6.45×10^−3^; log_2_FC_PHG_=0.43, p_PHG_=4.40×10^−4^), but not between male AD cases and controls (log_2_FC_DLPFC_=6.86×10^−3^, p_DLPFC_=0.98; log_2_FC_PHG_=0.22, p_PHG_=0.18). Second, *DCHS2* is differentially expressed in the temporal cortex (TCX) between male AD cases and controls (log_2_FC_TCX_=-0.31, p_TCX_=4.65×10^−2^), but not between female AD cases and controls (log_2_FC_TCX_=-0.22, p_TCX_=0.15). *TRIM2* likewise is differentially expressed in TCX between male AD cases and controls (log_2_FC_TCX_=-0.24, p_TCX_=0.02), but not between female AD cases and controls (log_2_FC_TCX_=-0.09, p_TCX_=0.33). Third, *AGA* is differentially expressed in the Cerebellum (CBE) between male AD cases and controls (log2FC_CBE_=-0.57, p_CBE_=1.48×10^−3^), but not between female AD cases and controls (log2FC_CBE_=-0.18, p_CBE_=0.42). Finally, *SAR1A* is differentially expressed in the CBE between male AD cases and controls (log_2_FC_CBE_=-0.11, p_CBE_=0.04), but not between female AD cases and controls (log_2_FC_CBE_=-0.06, p_CBE_=0.34).

Lastly, we leveraged our gene-level analysis (with MAGMA; see above) to evaluate whether or not established AD risk genes with evidence of sex specificity (*BIN1*^25,26^, *SORL1*^27,28^, *PICALM*^29^, *MS4A6A*^26^, and *SLC9A7*^30^) were associated with any of the 3 cognitive domains. For this manuscript, we did not evaluate the *APOE* locus, as our group has published an extensive sex-specific study on the relationship of polymorphism at the *APOE* locus and cognitive endophenotypes^31^. As shown in **Table 3**, all genes (*BIN1*^25,26^, *PICALM*^29^, *MS4A6A*^26^, and *SLC9A7*^30^) displayed evidence of sex-specificity with cognitive endophenotypes, except for *SORL1*. Two patterns that emerged through this analysis were that several AD risk genes were significantly (p<0.05) associated with baseline executive functioning in females only and with language decline in males only (**Table 3**). Notably, *PICALM* was significantly associated in both sexes and across multiple domains, and prior published evidence suggests that sex modifies the relationship of *PICALM* with disease processes, including neuroimmunity^29^.

**Table 3.**
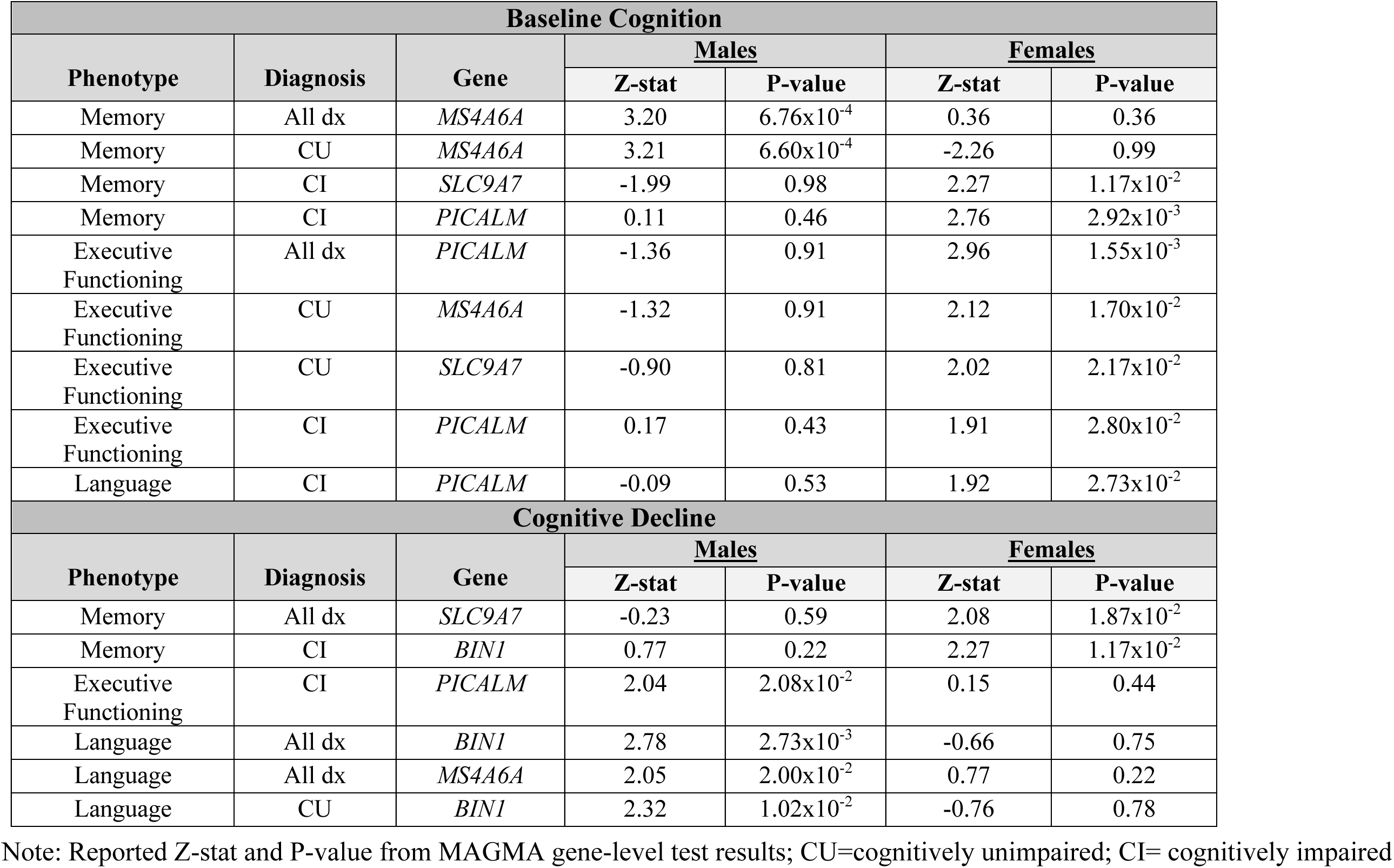
Significant Results from Sex-Specific AD Candidate Gene Analysis.

### Whole-Genome Results including Pathway Analyses and Genetic Covariances

We conducted pathway analyses with MAGMA (see **Methods**), and results are presented in **Supplementary Tables 13-18.** We identified multiple sex-specific pathways, defined as pathways that survived adjustment for multiple comparisons (FDR<0.05) in at least one sex or had an FDR-significant sex-interaction. Overall, patterns emerged that pathways significantly relating to cognitive endophenotypes in a sex-specific manner fell into four overarching groups: (1) DNA and polypeptide formation, (2) Cell division and morphology, (3) Metabolic process, and (4) Extracellular matrix activity.

We conducted a proof-of-principle genetic covariance test (GNOVA^32^) with a well-established sex-agnostic educational attainment GWAS^33^ to see if there was evidence at the whole-genome level for a genetic association between education and cognition, by sex. As expected, we identified significant shared genetic architecture between genetic predisposition for higher educational attainment and better baseline cognition across all 3 domains in males (p_memory_=1.60×10^−3^; p_executive_functioning_=2.75×10^−14^; p_language_=1.18×10^−4^) and in females (p_memory_=3.35×10^−22^; p_executive_functioning_=1.74×10^−25^; p_language_=3.33×10^−15^). Furthermore, as expected, we identified significant shared genetic architecture between genetic predisposition for higher educational attainment and slower cognitive decline across 2 domains in males, with the third domain just falling below significance (p_memory_=5.51×10^−3^; p_executive_functioning_=5.08×10^−4^; p_language_=0.09) and 3 domains in females (p_memory_=1.17×10^−5^; p_executive_functioning_=2.77×10^−3^; p_language_=1.38×10^−6^). Associations trended more significant in females compared to males when evaluating both baseline cognition and longitudinal cognitive decline.

We additionally conducted sex-specific genetic covariance analyses (GNOVA^32^) by leveraging sex-stratified GWAS summary statistics from the UKBB^23^. Specifically, we evaluated a curated set of male and female sex hormone-related traits that are known to effect testosterone or estrogen exposure, to determine if there was shared genetic architecture with cognition. Prior GWAS on circulating hormone levels^34,35^ and sex-agnostic genetic correlation results from our group on traits affecting circulating estrogen levels^19^ suggest preliminary evidence of a sex-specific genetic basis to circulating sex hormone levels. In total, we tested 29 traits (8 male traits, 21 female traits). Within each phenotype, we adjusted for multiple comparisons across the 8 male traits and the 21 female traits, separately, and we reported all genetic covariances with an FDR-corrected p<0.05 in **Table 4**.

**Table 4.** Significant Genetic Covariances between Cognition and Hormone-Related Traits.

| **Baseline Cognition** |  |  |  |  |  |
| --- | --- | --- | --- | --- | --- |
| **UKBB Trait** | **UKBB Code** | **Phenotype** | **Sex** | **Rho** | **P.FDR** |
| Enlarged Prostate | 20002_1396 | Executive Functioning | Men | -0.02 | $9.11 \times 10^{-3}$ |
| Age began oral contraceptive pill | 2784 | Memory | Women | -0.02 | $1.68 \times 10^{-4}$ |
| Miscarriage | 20002_1559 | Memory | Women | -0.01 | $7.72 \times 10^{-4}$ |
| Age at menarche | 2714 | Memory | Women | -0.02 | $1.71 \times 10^{-2}$ |
| Length of menstrual cycle | 3710 | Memory | Women | -0.03 | $1.71 \times 10^{-2}$ |
| Age at menarche | 2714 | Executive Functioning | Women | -0.02 | $2.44 \times 10^{-2}$ |
| Every taken HRT | 2814 | Executive Functioning | Women | 0.01 | $4.28 \times 10^{-2}$ |
| Age began oral contraceptive pill | 2784 | Executive Functioning | Women | 0.01 | $4.70 \times 10^{-2}$ |
| Menorrhagia | 20002_1556 | Language | Women | -0.02 | $1.24 \times 10^{-3}$ |
| Female infertility | 20002_1403 | Language | Women | 0.01 | $3.91 \times 10^{-3}$ |
| **Cognitive Decline** |  |  |  |  |  |
| **UKBB Trait** | **UKBB Code** | **Phenotype** | **Sex** | **Rho** | **P.FDR** |
| Testicular problems | 20002_1214 | Language | Men | -0.02 | $9.28 \times 10^{-3}$ |
| Length of menstrual cycle | 3710 | Memory | Women | -0.03 | $2.91 \times 10^{-2}$ |
| Had menopause | 2724 | Memory | Women | 0.01 | $2.91 \times 10^{-2}$ |
| Number of live births | 2734 | Memory | Women | 0.01 | $2.91 \times 10^{-2}$ |
| Number of miscarriages | 3839 | Executive Functioning | Women | -0.03 | $2.27 \times 10^{-3}$ |
| Had hysterectomy | 3591 | Executive Functioning | Women | 0.02 | $7.27 \times 10^{-3}$ |
| Had menopause | 2724 | Executive Functioning | Women | 0.02 | $2.39 \times 10^{-2}$ |
| Uterine fibroids | 20002_1351 | Executive Functioning | Women | 0.01 | $2.39 \times 10^{-2}$ |
| Uterine fibroids | 20002_1351 | Language | Women | 0.02 | $5.65 \times 10^{-3}$ |

Note: Genetic covariances (Rho) reported from GNOVA software tool; P-values adjusted for multiple comparisons (i.e., false-discovery rate - FDR); UKBB=UK Biobank

Among males, we identified significant genetic covariances between cognition and traits influencing testosterone exposure. First, we identified a significant genetic covariance with baseline cognition (FDR<0.05), including identification of shared genetic architecture between “enlarged prostate” and baseline executive functioning (ρ=-0.02, P.FDR=9.11×10^−3^). Additionally, genetic predisposition to “testicular problems” was significantly associated with the genetic architecture of language decline (ρ=-0.02, P.FDR=9.28×10^−3^).

Among females, we identified significant genetic covariances between cognition and traits influencing estrogen exposure. First, we identified 9 traits with significant genetic covariances with baseline cognition (FDR<0.05). We identified 4 traits genetically associated with baseline memory, including a top association, “age began oral contraceptive pill” (ρ=-0.02, P.FDR=1.68×10^−4^). We identified 3 traits genetically associated with baseline executive functioning, including a top association, “age at menarche” (ρ=-0.02, P.FDR=2.44×10^−2^). We identified 2 traits genetically associated with baseline language, including top association, “menorrhagia” (ρ=-0.02, P.FDR=1.24×10^−3^). Next, we identified 8 traits with significant genetic covariances with cognitive decline (FDR<0.05). We identified 3 traits genetically associated with memory decline, including a top association “having had menopause” (ρ=-0.03, P.FDR=2.91×10^−3^). We identified 4 traits genetically associated with executive functioning decline, including top association, “number of miscarriages” (ρ=-0.03, P.FDR=2.27×10^−3^). Finally, we identified one trait genetically associated with language decline, which was “uterine fibroids” (ρ=0.02, P.FDR=5.65×10^−3^).

## Discussion

We conducted a large-scale, cross-ancestral meta-analysis to identify genetic factors associated with memory, executive functioning, and language in a sex-specific manner. While the heritability of each cognitive domain was comparable across sexes, we identified 3 sex-specific genomic loci, including a chromosome 2 locus, *VRK2* (rs13387871; **Figure 1**; **Supplementary Figure 1**) associated with female-specific language decline, a chromosome 4 locus, *DCHS2*, (rs12501200; **Figure 2**; **Supplementary Figure 2**) associated with memory decline in CI males, and a sex-interaction with baseline executive functioning among CI individuals at *AGA* (rs1380012; **Figure 3**; **Supplementary Figure 3**). We also present evidence for shared genetic architecture between traits influencing lifetime estrogen exposure and AD-related cognitive decline. Overall, our study shows that at the variant, gene, and whole-genome levels, there is a sex-specific genetic basis for cognitive decline in older adults.

Based on our sex-specific meta-analysis, brain eQTL evidence, and differentially expression findings in brain, we identified the *VRK2/FANCL* gene cluster (led by rs13387871; **Figure 1**; **Supplementary Figure 1**) as a candidate gene of interest for female-specific language decline. *VRK2* has been previously implicated as a candidate gene across a host of neuropsychiatric and neurological traits, including schizophrenia^36,37^, multiple sclerosis (MS)^37^, bipolar disorder^38,39^, major depressive disorder^36^, circadian rhythm^40,41^, insomnia^42^, musical beat synchrony^43^, and developmental stuttering^44^. Additionally, *VRK2* has shown some sex-specificity in the literature, as *VRK2* gene expression was downregulated in women with MS 40+ years of age (compared to healthy controls)^37^, and in a prior sex-specific GWAS study of our group’s, we identified female-specific shared genetic architecture between MS and cognitive resiliency^20^. Furthermore, *VRK2* has shown a male-specific GWAS association for higher odds of developmental stuttering^44^. In our study, the minor allele of rs13387871 (*VRK2*) harbored protection against language decline in females, which would complement the MS finding but differ from the stuttering finding. It may be the case that *VRK2* exhibits one type of sex-specific role in neurodevelopment, but perhaps during aging *VRK2* may switch roles, playing a different sex-specific role in neurodegeneration. In addition, *VRK2* was the top locus in a musical beat synchrony GWAS^43^. Musical beat synchrony is a language-related trait, and notably developmental stuttering^44^ and schizophrenia^45^ are also traits with a significant language component. Taken together with our findings, perhaps *VRK2* plays a sex-specific role in language ability, but in a differing manner in neurodevelopment vs. disease. Future work should continue to evaluate the sex-specific role of the *VRK2* locus in neurodevelopment, aging, and neurodegenerative disease, particularly as this gene relates to language performance.

We identified numerous other candidate genes that relate to cognitive endophenotypes in a sex-specific manner. One of these was led by an association with rs12501200 (**Figure 2**; **Supplementary Figure 2**) and memory decline among CI males. In our previous sex-specific genetic analysis of late-life memory performance^18^, rs12501200 reached suggestive significance (p<1×10^−5^) but not genome-wide significance (p<5×10^−8^). In this current study, with additional samples, this locus surpassed the stringent genome-wide significance threshold (p<5×10^−8^), confirming the importance of the locus, led by rs12501200, in rate of memory decline among CI males. The lead variant in the locus is closest to *DCHS2* and shows brain eQTL evidence as well as male differential expression in brain for this gene. *DCHS2* is a previously reported AD gene, associated with Braak neurofibrillary tangle (NFT) stage^46^, interacting with COPS5 that interfaces with APP, and additionally *DCHS2* is associated with AD age-at-onset^47^. *DCHS2* also shows some sex-specificity the literature, with male-specific associations with skeletal strength indices related to osteoporosis^48^. In addition, we identified two more candidate genes through gene-set analyses that have been previously published in relation to AD and are significantly associated with memory decline among CI males in our study. The first gene, *TNN*, has been previously associated with pathologically-confirmed, rapidly progressing AD^49^, and the second gene, *SAR1A,* is widely expressed in the brain, is downregulated in the AD brain, and is associated with amyloid plaque load^50^.

In a prior study of our group on the sex-specific genetic study of cognitive resilience, we discussed the possible cross-talk between genetic factors, cognition, and circulating sex hormones, specifically estrogen. We add more evidence from the present study to support that hypothesis. First, in our candidate approach evaluating known AD risk genes, we identified sex-specificity at a recently discovered X-chromosome AD locus, *SLC9A7*^30^, that was significantly associated with higher odds of AD and appeared to be driven by males^30^. In our study, a candidate AD gene approach identified nominal significance for *SLC9A7* in females (**Table 3**; p<0.05, gene-level tests), associated with baseline memory and executive functioning and rate of memory decline, complementing the sex-specific findings in Belloy and colleagues recent study^30^. Furthermore, in our genetic correlation analysis with female and male hormone-related traits, we identified shared genetic architecture between estrogen-related traits and cognitive endophenotypes in females. Multiple studies show relationships between circulating estrogen levels and cognitive decline, suggesting that features promoting higher and/or longer estrogen exposure throughout life may be neuroprotective against cognitive decline^51–53^. In our study, we provide some evidence for a genetic basis for this, showing that pathways of relevance to genetic susceptibility of both female estrogen-related traits and AD-related cognition may, in part, be shared. The relationship between the X-chromosome, lifetime estrogen exposure, and AD-related cognitive decline is a growing area of interest in the field. We feel we provide exciting preliminary evidence in support of the current literature and further demonstrate that there may be a sex-specific genetic basis to the cross-talk between lifetime estrogen exposure and multiple domains of cognition in older females.

Our study had many strengths. We leveraged high-quality, standardized and harmonized cognitive scores from the ADSP-PHC, performed cross-ancestry analyses among NHW and NHB individuals, and included the X-chromosome, which is often left out of common variant AD genetic studies. However, our study is not without limitations. We were unable to replicate 2 of the 3 sex-specific loci identified in this study. Additionally, we had an imbalance of NHW and NHB individuals, the latter including far fewer individuals. Our cross-ancestry analysis was also restricted to NHW and NHB individuals, limiting the generalizability of our findings to other ancestry groups. Further, many individuals in our analysis had comorbid etiologies, which may have influenced our findings, and future work should delve into the interplay of multiple etiology AD and cognition in older adults.

Overall, we identified multiple sex-specific genetic loci, candidate genes, and biological pathways associated with memory, executive functioning, and language performance, all of which we demonstrated are heritable traits in both sexes. Our findings contribute to the understanding of the genetic basis for preclinical and clinical AD by sex.

## Methods

### Study Participants

Please see participant characteristics presented in **Table 1**. Our discovery sample included 33,918 participants, consisting of 14,539 (42.9%) males and 19,379 (57.1%) females. Of these participants, 10,898 (75.6%) males and 14,444 (74.5%) females had data for 2 or more visits. Participants spanned 10 cohorts of aging and AD, with 9 of these cohorts containing data across multiple visits. The 10 cohorts included in this study were: the Anti-Amyloid Treatment in Asymptomatic Alzheimer’s Disease (A4; cross-sectional study)^54^, Adult Changes in Thought (ACT)^55^, Alzheimer’s Disease Neuroimaging Initiative (ADNI; https://adni.loni.usc.edu), Biomarkers of Cognitive Decline Among Normal Individuals (BIOCARD)^56^, Baltimore Longitudinal Study of Aging (BLSA)^57^, Knight Alzheimer’s Disease Research Center (Knight ADRC)^58^, National Alzheimer’s Coordinating Center (NACC)^59–63^, National Institute on Aging Alzheimer’s Disease Family Based Study (NIA-AD FBS)^64^, Religious Orders Study/Memory and Aging Project/Minority Aging Research Study (ROS/MAP/MARS)^65,66^, and the Wisconsin Registry for Alzheimer’s Prevention (WRAP)^67^. Please see **Supplementary Methods I** for an overview of each cohort study. Participants from all cohorts completed written informed consent, and all research was conducted in accordance with each cohort’s Institutional Review Board-approved protocols. Secondary analyses of all data were approved by the Vanderbilt University Medical Center Institutional Review Board.

### Clinical Diagnosis Determination

Each of the 10 cohorts included in our study had their own protocols for clinical diagnosis determination and those protocols can be found in each cohort’s published papers. In general, all studies followed the National Institute of Neurological and Communicative Disorders and Stroke and the Alzheimer’s Disease and Related Disorders Association (NINCDS-ADRDA) standardized criteria. For the purposes of this study, we desired to have a harmonized diagnosis variable across all cohorts to facilitate conducting secondary analyses by diagnostic subgroup. We assigned study-specific diagnostic codes to (1) cognitively unimpaired, (2) mild cognitive impairment (MCI), or (3) clinical dementia due to Alzheimer’s disease (AD). Because of the variability in sample size of MCI-diagnosed participants across studies, we created a diagnostic binary classifier of either cognitively unimpaired (CU) or cognitively impaired (CI), whereby the former included all CU individuals, and the latter included those with an MCI or AD dementia clinical diagnosis. To maximize sample size, individuals with multiple etiologies, such as MCI or AD with other neuropathologies or comorbid conditions were left in the sample.

### Genetic Quality Control and Imputation

We applied an in-house standardized quality control (QC) and imputation pipeline to raw genetic data from each cohort. For information regarding genotyping arrays used by each cohort, please see the **Supplementary Methods II.A**. Variant-level QC included filtering variants with >5% missingness, <1% minor allele frequency (MAF), and separating data into autosomes and X-chromosome (please see **Supplementary Methods II.B.** for additional X-chromosome-specific processing steps). Sample-level QC included filtering samples with >1% missingness, samples with excessive relatedness (pi-hat>0.25), samples with mismatched self-reported and genetically determined sex, and samples with excessive heterozygosity. Self-reported race and ethnicity and a principal component analysis (PCA) were leveraged to assign genetic ancestry, the latter of which evaluated genetic similarity to 1000 Genomes reference super populations. The rest of the QC was performed in two datasets, one including samples assigned to non-Hispanic white (NHW) race and ethnicity, and another dataset of samples spanning all race and ethnicity groups. All QC steps to follow were performed separately in the NHW and all race datasets. A Hardy-Weinberg Equilibrium (HWE) exact test was performed next, removing variants that were not in accordance with HWE (p<1×10^−6^), data were lifted to GRCh38 (variants failing liftover removed), and alleles mismatching with the Trans-Omics for Precision Medicine (TOPMed) server were removed^68–70^. Cleaned genetic data were phased and imputed on the TOPMed^68–70^ imputation server, and poor quality variants with an imputed R^2^<0.8 as well as duplicated and multi-allelic variants, and variants with a MAF<1% were removed post-imputation. Finally, for the scope of this analyses we desired to have a NHW and a non-Hispanic black (NHB) dataset, the former of which we already had and the latter of which we obtained via a combination of PCA, comparing similarity to 1000G AFR, and self-reported race and ethnicity. At this stage, for applicable datasets, we implemented a procedure for merging across chip, which is described in **Supplementary Methods II.C.** Lastly, we performed an iterative outlier removal procedure on the imputed, cleaned NHW and NHB datasets to obtain the final, cleaned, imputed genetic data. Further details regarding our in-house QC and imputation protocol are available upon request.

### Cognitive Harmonization

The ADSP-PHC harmonized cognitive data across 10 cohorts of aging and AD to facilitate joint analysis. Our group has previously published this harmonization protocol^16^ and have successfully leveraged the resulting data for multiple published genetic studies^17,18,31^. We will overview the harmonization protocol to follow. Qualified neuropsychologists and behavioral neurologists (authors A.J.S., J.M., and E.H.T.) took each individual item from neuropsychological test batteries and assigned items to cognitive domains, including memory, executive functioning, language, visuospatial ability, or none of the above. To ensure that items across studies had similar meanings, test items were recoded to match one another such that +1 point on a given test item meant exactly 1-point better performance in each study. The next step was to determine which model best fits the data and to do this, the most recent visit was selected for each participant and placed into a confirmatory factors analysis (CFA) whereby the best single-factor or bi-factor model was chosen. Fit statistics (i.e., confirmatory fit index, Tucker-Lewis Index, and root mean square error of approximation) were used to choose the best fit model. Then to facilitate co-calibration, anchor items were selected, defined as shared test items across studies, and these items were placed into their own model to retrieve a set of parameters. A full CFA with all datapoints was conducted, whereby anchor items were given set, shared weights and non-shared test items were allowed to be included in the model and freely estimated. The final factor scores were extracted from the model, which resulted in cognitive composite scores on the same scale across all 10 cohorts.

### Cognitive Endophenotypes

We generated 6 cognitive endophenotypes from the harmonized cognitive scores, 3 baseline cognition endophenotypes and 3 cognitive decline endophenotypes. Baseline cognition included memory, executive functioning, and language and were generated all the same across all cohorts and participants (note – A4 was a cross-sectional study and solely had memory scores). To gather baseline cognition, we selected the cognitive score from each participant’s first visit for each respective cognitive domain. To generate measures of cognitive decline, we calculated cognitive slopes for each participant for each cognitive domain. Slopes were calculated with a linear mixed effects model, allowing both slope and intercept to vary per individual. Inclusion into the model required an individual to have scores for 2+ timepoints for a given cognitive domain, and all available datapoints were leveraged in each model. As a result, we had generated 3 cognitive slopes per individual to represent rates of decline for memory, executive functioning, and language, respectively.

### Statistical Analyses

#### SNP-Based Heritability Estimates

We calculated SNP-based heritability estimates with the Genome-wide Complex Trait Analysis (GCTA) software package^71^. This package allows for leveraging genotype-level data, instead of summary statistics, to calculate heritability, typically allowing for more stable estimates. To calculate heritability, we calculated genetic relationship matrices (GRM) among males and females separately, and within each ancestry group. Then we fed the GRM and the cognitive endophenotypes for each sample into a restricted maximum likelihood (REML) model to result in SNP-based heritability estimates per sample. To determine if heritability differed by sex, we implemented a published algorithm^21^.

#### Ancestry-Specific Genome-Wide Association Studies

Prior to conducting genome-wide association studies (GWAS), we retained individuals with a baseline age of ≥50 years old, and we assessed cryptic relatedness across all 10 cohorts, filtering one sample per related pair. We began with individual cohort GWAS and restricted each cohort to first NHW and then to NHB ancestry groups. Cohort GWAS for each ancestry group were conducted if the sample size was ≥50 (for NHB this resulted in BLSA, NACC, and ROS/MAP/MARS only). In total, we had 6 different GWAS outcomes, which included 3 cognitive endophenotypes at baseline – baseline memory, baseline executive functioning, and baseline language, as well as 3 endophenotypes of cognitive decline – memory decline, executive functioning decline, and language decline. For each cohort and each ancestry group, and within each GWAS outcome, we performed 3 GWAS: male, female, and sex-interaction. Both the sex-stratified and sex-interaction GWAS were conducted with PLINK^72^ (v1.9b) using the linear association model with additive variant coding. Models were adjusted for age at baseline visit and 5 genetic ancestry principal components. Additionally, the sex-interaction GWAS included a sex*SNP interaction term of interest. We performed two more sets of identical models, first restricting the sample to CU individuals and then to CI individuals. In addition, we conducted an identical set of models for the X-chromosome, except male genotypes were coded as 0/2, while female genotypes were still coded as 0/1/2, to account for X-chromosome dosage differences between sexes.

#### Ancestry-Specific Genome-Wide Meta-Analysis

We next performed genome-wide meta-analyses across cohorts and for each ancestry group (i.e., NHW and NHB) among males, females, and with a sex-interaction. Secondary meta-analyses were additionally performed with identical models with the sample first restricted to CU and then to CI. Prior to analysis, we retained variants from each GWAS if the minor allele count for a given variant was ≥10 for a given stratum-specific contingency table (to ensure we retained common variants). Meta-analyses were performed with a fixed-effects model (i.e., GWAMA v. 2.2.2)^73^. Variants were filtered out post-hoc if absent from >2 cohorts for NHW-specific meta-analyses and >1 cohort for NHB-specific meta-analyses.

#### Cross-Ancestry Genome-Wide Meta-Analysis

In addition to ancestry-specific analyses, we were interested in effects across ancestry groups, and to do this, we performed cross-ancestry meta-analyses (fixed-effects model; GWAMA v. 2.2.2)^73^. Variants were retained if present in both ancestry groups (i.e., NHW and NHB) to ensure we were detecting cross-ancestral SNP effects. Secondary analyses were conducted among CU and CI diagnosis subgroups.

#### Sensitivity Analysis

Educational attainment, cognition, and AD have a well-established relationship. More specifically, higher educational attainment has been linked to a lower risk of MCI/AD^74,75^ and to better cognition across multiple domains, including memory, executive functioning, attention, and verbal fluency, even when adjusting for AD risk factors^74^. Evidence shows a shared genetic architecture between cognitive performance, AD, and educational attainment^19,75^. Thus, we conducted a set of sensitivity analyses wherein we used an identical set of models as our main analysis with the addition of adjusting for years of education, as a proxy for education attainment. We took this step to clarify whether any part of genome-wide signals identified may, in part, be due to the influence of education on AD-related cognitive performance.

#### Replication Analysis

We used two replication cohorts to attempt replication on genome-wide significant cross-ancestry loci. First, we leveraged a publicly available, open-access database from the UK Biobank (UKBB)^23^ containing thousands of summary statistics of sex-stratified GWAS performed on health and medical traits. Specifically, we attempted replication in sex-stratified traits related to cognitive performance or family history of AD. Second, we sought replication in an independent cohort, the Framingham Heart Study (FHS^22^), which was originally designed as an epidemiological study to investigate cardiovascular traits^76^. Replication models conducted in FHS tested the association of any genome-wide significant loci found in our discovery cohort with similarly-derived harmonized cognitive endophenotypes^16,22^. For both methods of replication, significance was set at p<0.05.

#### Functional Annotation of Genome-Wide Significant Loci

We leveraged a series of publicly available databases to find functional evidence that may explain the biological relevance underlying genome-wide significant loci. We queried index variants in the following quantitative trait loci (QTL) databases: xQTLatlas (http://www.hitxqtl.org.cn/), GTEx (https://gtexportal.org/), BrainSeq (http://eqtl.brainseq.org/phase2/eqtl/), and eQTLGen Consortium (whole blood; https://www.eqtlgen.org). We queried genes prioritized from the prior steps in combination with significant gene-level test results (see next section) in AGORA, a platform containing evidence for associations with AD across omics layers (https://agora.adknowledgeportal.org). One additional measure leveraged was evaluating the association of established AD risk genes, with published sex-specificity, to determine if any known AD risk genes were associated with baseline cognition or cognitive decline in a sex-specific manner. We deemed an association significant in this candidate approach with p<0.05.

#### Gene- and Pathway-Level Analyses

We performed gene-set analyses with the Multi-marker Analysis of GenoMic Annotation (MAGMA^24^ v.1.09) software. Prior to analysis, we removed a 1 Mb region up- and down-stream of the well-characterized *APOE* locus. To annotate SNPs to genes, we leveraged NCBI b38 gene annotations and SNP information from our genetic data files. Creation of pathway annotation files included a compilation of pathway annotations from MSigDB^77^, which included annotation sets from Gene Ontology (GO), Kyto Encyclopedia of Genes and Genomes (KEGG), Reactome DB, BioCarta, and Pathway Interaction Database (PID). Gene tests were conducted first, leveraging cross-ancestry meta-analysis results, and tests were performed to determine if more SNPs were significantly associated than expected by chance, within a given gene window, with each cognitive endophenotype. A similar style test was next conducted for pathway-level tests, whereby the results of the gene-level tests were leveraged to conduct these pathway tests. Both gene and pathway tests were conducted in a sex-stratified manner and with a sex-interaction. All tests were adjusted for multiple comparisons with the false discovery rate (FDR) procedure, with significance set at FDR<0.05.

#### Genetic Covariance Analysis

We evaluated if there was shared genetic architecture between cognition and a curated set of complex traits, in a sex-specific manner. To conduct this analysis, we leveraged a set of sex-stratified GWAS of health and medical traits from the UKBB^23^. These publicly available GWAS can be downloaded from the Neale lab website (http://www.nealelab.is/uk-biobank)^23^. Due to the UKBB GWAS consisting of European samples, we matched on ancestry using our NHW ancestry-specific meta-analyses (with the *APOE* locus removed). To conduct genetic covariance tests, we used the Genetic Covariance Analyzer (GNOVA)^32^ software, which our group has leveraged prior to identify genetic covariances, including by sex^18,20^. Prior to calculating genetic covariance estimates, every SNP effect from each GWAS was converted to be on a z-score scale. Then we proceeded to calculate genetic covariances between trait pairs, and GNOVA corrects for LD-inflation with ancestry-matched LD-scores (i.e., 1000 Genomes EUR reference panel). For the scope of this analysis, we analyzed a subset of traits (8 male, 21 female) that were male- or female-specific hormone-related traits (i.e., estrogen and testosterone), and we adjusted for multiple comparisons within each phenotype and sex (FDR<0.05). For those interested in results beyond the curated set of traits presented in this manuscript, please contact corresponding author, Dr. Dumitrescu.

## Data Availability

All GWAS summary statistics generated from this analysis are publicly accessible on the National Institute on Aging Genetics of Alzheimer’s Disease Data Storage Site (NIAGADS) website: https://dss.niagads.org/. Harmonized cognitive scores leveraged in this analysis are available as part of the ADSP available on https://dss.niagads.org/. A description of all available phenotypes in the ADSP-PHC is listed online at https://www.vmacdata.org/adsp-phc.

## Code Availability

Code for this project is available upon request. Please send all requests to corresponding author, Dr. Dumitrescu at.

## Acknowledgements

The ADSP Phenotype Harmonization Consortium (ADSP-PHC) is funded by NIA (U24 AG074855, U01 AG068057 and R01 AG059716). The harmonized cohorts within the ADSP-PHC include: the Anti-Amyloid Treatment in Asymptomatic Alzheimer’s study (A4 Study), a secondary prevention trial in preclinical Alzheimer’s disease, aiming to slow cognitive decline associated with brain amyloid accumulation in clinically normal older individuals. The A4 Study is funded by a public-private-philanthropic partnership, including funding from the National Institutes of Health-National Institute on Aging, Eli Lilly and Company, Alzheimer’s Association, Accelerating Medicines Partnership, GHR Foundation, an anonymous foundation and additional private donors, with in-kind support from Avid and Cogstate. The companion observational Longitudinal Evaluation of Amyloid Risk and Neurodegeneration (LEARN) Study is funded by the Alzheimer’s Association and GHR Foundation. The A4 and LEARN Studies are led by Dr. Reisa Sperling at Brigham and Women’s Hospital, Harvard Medical School and Dr. Paul Aisen at the Alzheimer’s Therapeutic Research Institute (ATRI), University of Southern California. The A4 and LEARN Studies are coordinated by ATRI at the University of Southern California, and the data are made available through the Laboratory for Neuro Imaging at the University of Southern California. The participants screening for the A4 Study provided permission to share their de-identified data in order to advance the quest to find a successful treatment for Alzheimer’s disease. We would like to acknowledge the dedication of all the participants, the site personnel, and all of the partnership team members who continue to make the A4 and LEARN Studies possible. The complete A4 Study Team list is available on: a4study.org/a4-study-team.; the Adult Changes in Thought study (ACT), U01 AG006781, U19 AG066567; Alzheimer’s Disease Neuroimaging Initiative (ADNI): Data collection and sharing for this project was funded by the Alzheimer’s Disease Neuroimaging Initiative (ADNI) (National Institutes of Health Grant U01 AG024904) and DOD ADNI (Department of Defense award number W81XWH-12-2-0012). ADNI is funded by the National Institute on Aging, the National Institute of Biomedical Imaging and Bioengineering, and through generous contributions from the following: AbbVie, Alzheimer’s Association; Alzheimer’s Drug Discovery Foundation; Araclon Biotech; BioClinica, Inc.; Biogen; Bristol-Myers Squibb Company; CereSpir, Inc.; Cogstate; Eisai Inc.; Elan Pharmaceuticals, Inc.; Eli Lilly and Company; EuroImmun; F. Hoffmann-La Roche Ltd and its affiliated company Genentech, Inc.; Fujirebio; GE Healthcare; IXICO Ltd.;Janssen Alzheimer Immunotherapy Research & Development, LLC.; Johnson & Johnson Pharmaceutical Research & Development LLC.; Lumosity; Lundbeck; Merck & Co., Inc.;Meso Scale Diagnostics, LLC.; NeuroRx Research; Neurotrack Technologies; Novartis Pharmaceuticals Corporation; Pfizer Inc.; Piramal Imaging; Servier; Takeda Pharmaceutical Company; and Transition Therapeutics. The Canadian Institutes of Health Research is providing funds to support ADNI clinical sites in Canada. Private sector contributions are facilitated by the Foundation for the National Institutes of Health (www.fnih.org). The grantee organization is the Northern California Institute for Research and Education, and the study is coordinated by the Alzheimer’s Therapeutic Research Institute at the University of Southern California. ADNI data are disseminated by the Laboratory for Neuro Imaging at the University of Southern California; Estudio Familiar de Influencia Genetica en Alzheimer (EFIGA): 5R37AG015473, RF1AG015473, R56AG051876; the Health & Aging Brain Study – Health Disparities (HABS-HD), supported by the National Institute on Aging of the National Institutes of Health under Award Numbers R01AG054073, R01AG058533, R01AG070862, P41EB015922, and U19AG078109; the Korean Brain Aging Study for the Early Diagnosis and Prediction of Alzheimer’s disease (KBASE), which was supported by a grant from Ministry of Science, ICT and Future Planning (Grant No: NRF-2014M3C7A1046042); Memory & Aging Project at Knight Alzheimer’s Disease Research Center (MAP at Knight ADRC): The Memory and Aging Project at the Knight-ADRC (Knight-ADRC). This work was supported by the National Institutes of Health (NIH) grants R01AG064614, R01AG044546, RF1AG053303, RF1AG058501, U01AG058922 and R01AG064877 to Carlos Cruchaga. The recruitment and clinical characterization of research participants at Washington University was supported by NIH grants P30AG066444, P01AG03991, and P01AG026276. Data collection and sharing for this project was supported by NIH grants RF1AG054080, P30AG066462, R01AG064614 and U01AG052410. We thank the contributors who collected samples used in this study, as well as patients and their families, whose help and participation made this work possible. This work was supported by access to equipment made possible by the Hope Center for Neurological Disorders, the Neurogenomics and Informatics Center (NGI: https://neurogenomics.wustl.edu/) and the Departments of Neurology and Psychiatry at Washington University School of Medicine; National Alzheimer’s Coordinating Center (NACC): The NACC database is funded by NIA/NIH Grant U24 AG072122. SCAN is a multi-institutional project that was funded as a U24 grant (AG067418) by the National Institute on Aging in May 2020. Data collected by SCAN and shared by NACC are contributed by the NIA-funded ADRCs as follows: P30 AG062429 (PI James Brewer, MD, PhD), P30 AG066468 (PI Oscar Lopez, MD), P30 AG062421 (PI Bradley Hyman, MD, PhD), P30 AG066509 (PI Thomas Grabowski, MD), P30 AG066514 (PI Mary Sano, PhD), P30 AG066530 (PI Helena Chui, MD), P30 AG066507 (PI Marilyn Albert, PhD), P30 AG066444 (PI John Morris, MD), P30 AG066518 (PI Jeffrey Kaye, MD), P30 AG066512 (PI Thomas Wisniewski, MD), P30 AG066462 (PI Scott Small, MD), P30 AG072979 (PI David Wolk, MD), P30 AG072972 (PI Charles DeCarli, MD), P30 AG072976 (PI Andrew Saykin, PsyD), P30 AG072975 (PI David Bennett, MD), P30 AG072978 (PI Neil Kowall, MD), P30 AG072977 (PI Robert Vassar, PhD), P30 AG066519 (PI Frank LaFerla, PhD), P30 AG062677 (PI Ronald Petersen, MD, PhD), P30 AG079280 (PI Eric Reiman, MD), P30 AG062422 (PI Gil Rabinovici, MD), P30 AG066511 (PI Allan Levey, MD, PhD), P30 AG072946 (PI Linda Van Eldik, PhD), P30 AG062715 (PI Sanjay Asthana, MD, FRCP), P30 AG072973 (PI Russell Swerdlow, MD), P30 AG066506 (PI Todd Golde, MD, PhD), P30 AG066508 (PI Stephen Strittmatter, MD, PhD), P30 AG066515 (PI Victor Henderson, MD, MS), P30 AG072947 (PI Suzanne Craft, PhD), P30 AG072931 (PI Henry Paulson, MD, PhD), P30 AG066546 (PI Sudha Seshadri, MD), P20 AG068024 (PI Erik Roberson, MD, PhD), P20 AG068053 (PI Justin Miller, PhD), P20 AG068077 (PI Gary Rosenberg, MD), P20 AG068082 (PI Angela Jefferson, PhD), P30 AG072958 (PI Heather Whitson, MD), P30 AG072959 (PI James Leverenz, MD); National Institute on Aging Alzheimer’s Disease Family Based Study (NIA-AD FBS): U24 AG056270; Religious Orders Study (ROS): P30AG10161,R01AG15819, R01AG42210; Memory and Aging Project (MAP - Rush): R01AG017917, R01AG42210; Minority Aging Research Study (MARS): R01AG22018, R01AG42210; the Texas Alzheimer’s Research and Care Consortium (TARCC), funded by the Darrell K Royal Texas Alzheimer’s Initiative, directed by the Texas Council on Alzheimer’s Disease and Related Disorders; Washington Heights/Inwood Columbia Aging Project (WHICAP): RF1 AG054023;and Wisconsin Registry for Alzheimer’s Prevention (WRAP): R01AG027161 and R01AG054047. Additional acknowledgments include the National Institute on Aging Genetics of Alzheimer’s Disease Data Storage Site (NIAGADS, U24AG041689) at the University of Pennsylvania, funded by NIA. Important sources of funding for this study also included R01 AG073439 (PI, Logan Dumitrescu, MS, PhD) and F31 AG077791 (PI, Jaclyn M. Eissman, PhD).

## Author Information

### Consortia

**The Alzheimer’s Disease Neuroimaging Initiative (ADNI)**

Data used in preparation of this article were obtained from the Alzheimer’s Disease Neuroimaging Initiative (ADNI) database (adni.loni.usc.edu). As such, the investigators within the ADNI contributed to the design and implementation of ADNI and/or provided data but did not participate in analysis or writing of this report. A complete listing of ADNI investigators can be found at: http://adni.loni.usc.edu/wp-content/uploads/how_to_apply/ADNI_Acknowledgement_List.pdf

**The Alzheimer’s Disease Genetics Consortium (ADGC)**

**The Alzheimer’s Disease Sequencing Project (ADSP)**

