## Supplementary Methods & Figures for "Sex-Specific Genetic Drivers of Memory, Executive Functioning, and Language Performance in Older Adults"

### Supplementary Materials

#### Supplementary Methods

##### I. Overview of Included Datasets

Anti-Amyloid Treatment in Asymptomatic Alzheimer's Disease (A4): The A4 prevention trial[1] was created to test whether anti-amyloid therapeutics would help individuals with preclinical Alzheimer's disease (AD). This 3-year study enrolled 1,000 "at-risk" cognitively unimpaired adults 65 years of age or older. An individual was deemed to be at-risk for AD by presence of amyloid accumulation measured by an amyloid-PET scan. More about A4 can be found on their website: https://atri.usc.edu/study/a4-study/.

Adult Changes in Thought (ACT): The ACT study[2] was a prospective study that began in 1994, enrolling dementia-free participants 65 years of age or older. Enrollment focused on the Group Health Cooperative of Puget Sound (GHC) members in the Seattle area, which were comprised of a representative sample of the Seattle area surrounding GHC. The goal of this study was to track incident dementia cases, and participants had follow-up visits every 2 years. ACT successfully enrolled 2,581 participants at baseline with 2,356 participants that had >1 visit. More details of ACT can be found on their website: https://actagingresearch.org.

Alzheimer's Disease Neuroimaging Initiative (ADNI): The ADNI study is a multi-site, longitudinal study, with 4 completed phases that were included in our study, ADNI-1, ADNI-2, ADNI-GO, and ADNI-3 (and one phase currently being collected: ADNI-4). Overall, the goal of ADNI is to track and characterize progression from preclinical AD to severe AD dementia to better understand the sequence of AD biomarkers and their relation to cognitive changes. All ADNI data can be accessed at their non-embargoed website: http://adni.loni.usc.edu/.

Biomarkers of Cognitive Decline Among Normal Individuals (BIOCARD): The BIOCARD study[3] enrolled cognitively unimpaired middle-aged adults. The cohort was enriched for individuals with a family history of AD, whereby approximately 75% of individuals enrolled were first-degree relatives of someone with AD dementia. The cohort of adults were tracked over time to identify AD risk factors or biomarkers that were indicative of a transition to AD. More information about BICOARD can also be found on their website: http://www.biocard-se.org/.

Baltimore Longitudinal Study of Aging (BLSA): The BLSA study[4] is the biggest and longest-running longitudinal study of aging in America. Enrollment for BLSA began in 1958 and in total BLSA has enrolled over 3,000 cognitively healthy volunteers. The overarching goals of BLSA are to identify aging-related changes, including cognitive changes, physical changes, genetic factors, behavioral factors and environmental factors, as well as to study the relationship of aging with concomitant disease. For more information on BLSA please see their website: https://www.blsa.nih.gov.

Knight Alzheimer's Disease Research Center (Knight ADRC): The Knight ADRC[5] was founded in 1985 and aging research started being conducted in 1979, with the Memory and Aging Project as the first aging study. Inclusion criteria for Knight ADRC studies include being 40+ years of age, in good overall health, and dementia-free or mild dementia. Interval for participant follow-up on cognitive assessment is dependent on age. Over the past 30 years, the Knight ADRC has recruited greater than 6,600 individuals. More about the Knight ADRC data can be found here: https://dss.niagads.org/collections/knight-adrc-collection/.

National Alzheimer's Coordinating Center (NACC): NACC[6–10] is the hub of data collected from Alzheimer's Disease Research Centers (ADRCs), which began in 1999 when the National Institute on Aging established NACC. In total, NACC has collected data from more than 42 ADRCs and is currently collecting data from 42 of these ADRCs across 25 states. Over more than 20 years, NACC has obtained data on greater than 50,000 participants spanning the AD clinical spectrum, and 17,000 of these participants are still being followed to date. One important feature of NACC is that since 2005 all participating centers must implement the Uniform Data Set (UDS). The goal of the UDS is to obtain a standardized clinical dataset by providing each center with a standardized set of protocols for clinical assessment. Further information about NACC can be found on their website: https://naccdata.org.

National Institute on Aging Alzheimer's Disease Family Based Study (NIA-AD FBS): The NIA-AD FBS longitudinal study[11] began in 2003, with a focus on recruiting participants with AD or a family history of AD. Enrollment included older adults 60+ years of age who had at least two siblings with AD as well as age-matched healthy controls aged 55 or older. In total 1,756 families were recruited. NIA-AD FBS began implementing standardized cognitive assessment in 2006 and 75% of cohort members have partaken in this assessment. More about NIA-AD FBS can be found here: https://www.neurology.columbia.edu/research/research-centers-and-programs/national-institute-aging-alzheimers-disease-family-based-study-nia-ad-fbs.

Religious Orders Study/Memory and Aging Project/Minority Aging Research Study (ROS/MAP/MARS): Both ROS and MAP[12] are longitudinal clinical-pathological studies enrolling individuals across the AD clinical spectrum. ROS began in 1994 recruiting nuns, priests and brothers in the U.S., and MAP began in 1997 and recruited volunteers from the Chicago area. These studies collected a comprehensive host of data that included, genetic data, AD neuropathologic measures, resilience measures, measures of cognitive performance, and multiple layers of omics data. The MARS study[13] began in 2004, and is a longitudinal clinical-pathological study. Participants were recruited from the community in the Chicago area, and all enrolled participants were dementia-free African American individuals. More information on the ROS/MAP/MARS studies can be found on their website: https://www.radc.rush.edu.

Wisconsin Registry for Alzheimer's Prevention (WRAP): The WRAP study[14] is a longitudinal observational study that began in 2001. The goal of this cohort study is to identify midlife genetic and lifestyle factors that are associated with AD biomarkers and cognitive trajectories. Dementia-free participants between the ages of 40 and 65 (mean age 54) with a parental history of AD were enrolled in the study, with 1,561 total participants. All participants were followed 4 years after baseline and every 2 years to follow. More detailed information can be found on the WRAP website: https://wrap.wisc.edu.

#### II. Genetic Data Information

##### *A. Genotyping Array Information*

Here we list the genotyping array(s) leveraged for each cohort study:

Anti-Amyloid Treatment in Asymptomatic Alzheimer's Disease (A4) – Illumina Global Screening Array

Adult Changes in Thought (ACT) – Illumina Human660W-Quad Array

Alzheimer's Disease Neuroimaging Initiative (ADNI) – Illumina Human610-Quad BeadChip, Illumina HumanOmniExpress BeadChip, Illumina Omni 2.5M, and Illumina Global Screening Array v2

Biomarkers of Cognitive Decline Among Normal Individuals (BIOCARD) – obtained from the Alzheimer's Disease Genetics Consortium (ADGC)

Baltimore Longitudinal Study of Aging (BLSA) – Illumina NeuroChip and Illumina 550k

Knight Alzheimer's Disease Research Center (ADRC) – Illumina Human OmniExpress Beadchip, Illumina Human CoreExome Beadchip, Illumina Human 660k Quad Beadchip, Illumina NeuroX2, Illumina Human610-Quad Beadchip

National Alzheimer's Coordinating Center (NACC) – genetic data acquisition is described on their website

National Institute on Aging Alzheimer's Disease Family Based Study (NIA-AD FBS) – Illumina Human 610Quadv1_B Beadchip

Religious Orders Study/Memory and Aging Project/Minority Aging Research Study (ROS/MAP/MARS) – Global Screening Array-24 v3.0 BeadChip, Affymetrix GeneChip 6.0, and some data from ADGC GWAS datasets

Wisconsin Registry for Alzheimer's Prevention (WRAP) – Illumina Multi-Ethnic Genotyping Array

#### ***B. Additional Quality Control Steps for the X-Chromosome***

Due to the complexity of the X-chromosome, some additional steps were included in the quality control process which will be detailed to follow. To begin, the pseudo-autosomal (PAR) region was excluded. Variants were filtered by an additional test of differential missingness between sexes ($p<1x10^{-7}$). The Hardy-Weinberg Equilibrium Exact test ($p<1x10^{-6}$) was performed only in females, filtering variants out of the male sample accordingly.

#### ***C. Merging Across Genotype Chip***

Datasets requiring a merge across chip had additional quality control steps after final, clean, imputed genetic data by chip were ready. The following steps were conducted separately for the non-Hispanic white (NHW) and all races clean genetic data. First, we removed duplicated samples by first checking for concordance across the duplicated samples and then dropping the sample from both datasets if the concordance was <99% or from the less dense chip if the concordance was ≥99%. We next compared reference alleles across chips and dropped variants with reference allele mismatches. Additionally, we removed variants with minor allele frequency (MAF) differences across datasets greater than 10%. We merged across chip and then conducted a principal component analysis (PCA) to assess cryptic relatedness, performing an iterative outlier removal process to filter out any dataset outliers. The final, clean, imputed, and merged genetic data were now ready to use.

#### Supplementary Figures

##### Supplementary Figure 1. Forest Plot for Female-Specific Language Decline Locus Plotted by Cohort, Sex, and Ancestry

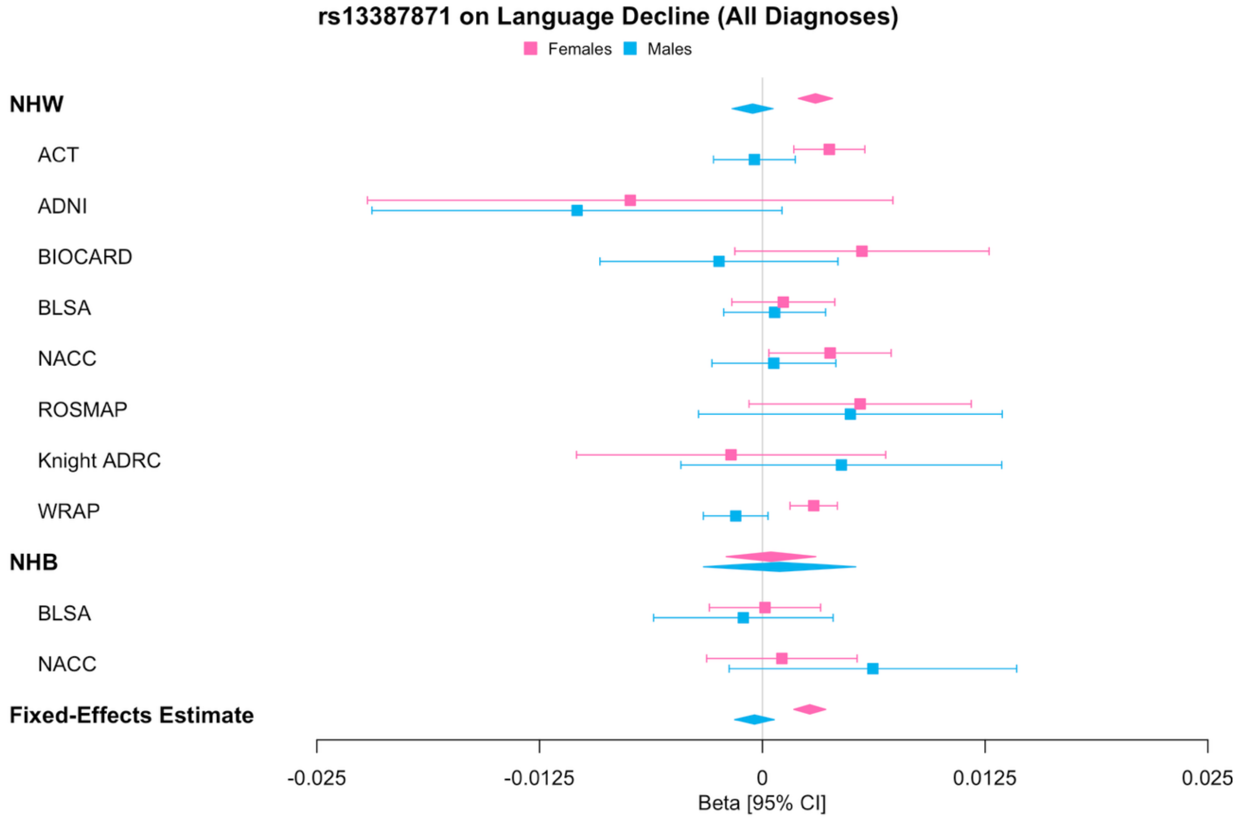


Note – NHW=non-Hispanic white; NHB=non-Hispanic black; cohort estimates are beta values from GWAS; overall ancestry-specific estimates are fixed-effects meta-analysis estimates, with the last fixed-effects estimate (bottom row) from the cross-ancestry meta-analysis

**Supplementary Figure 2. Forest Plot for Cognitively Impaired Male-Specific Memory Decline Locus Plotted by Cohort, Sex, and Ancestry**

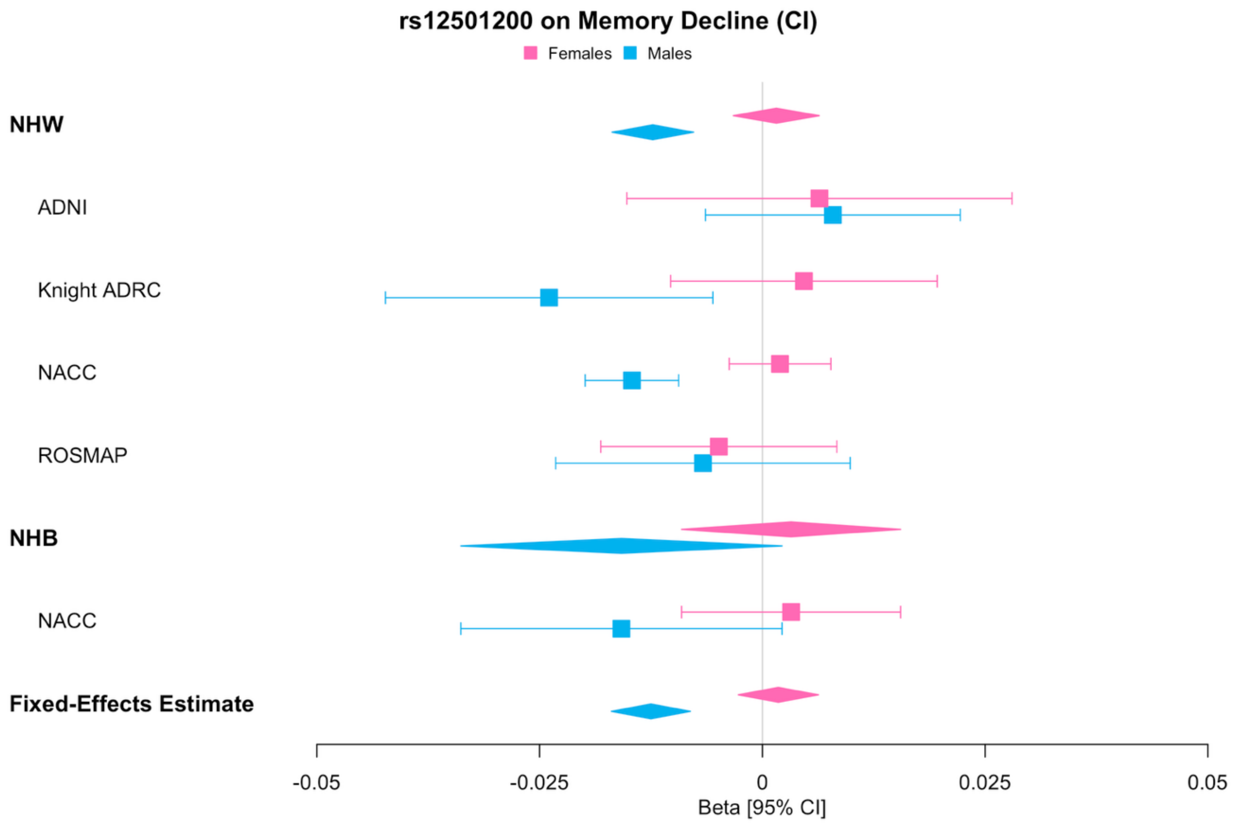


Note – NHW=non-Hispanic white; NHB=non-Hispanic black; CI=cognitive impaired (i.e., MCI or AD); cohort estimates are beta values from GWAS; overall ancestry-specific estimates are fixed-effects meta-analysis estimates, with the last fixed-effects estimate (bottom row) from the cross-ancestry meta-analysis

**Supplementary Figure 3. Forest Plot for Locus with Sex-Interaction with Baseline Executive Functioning among Cognitively Impaired Plotted by Cohort, Sex, and Ancestry**

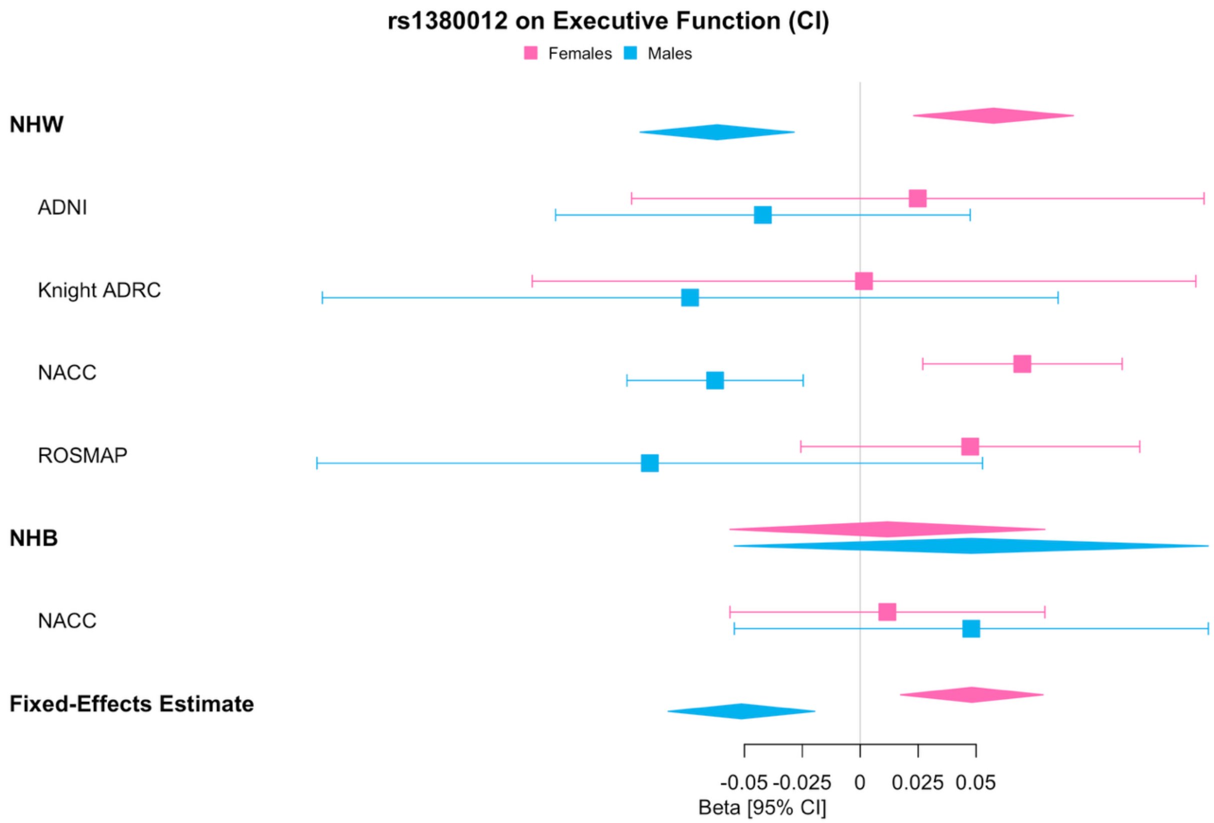


Note – NHW=non-Hispanic white; NHB=non-Hispanic black; CI=cognitive impaired (i.e., MCI or AD); cohort estimates are beta values from GWAS; overall ancestry-specific estimates are fixed-effects meta-analysis estimates, with the last fixed-effects estimate (bottom row) from the cross-ancestry meta-analysis
